# Predicting the need for electroconvulsive therapy via machine learning trained on electronic health record data

**DOI:** 10.1101/2025.06.19.25329678

**Authors:** Lasse Hansen, Jakob Grøhn Damgaard, Robert M. Lundin, Andreas Aalkjær Danielsen, Søren Dinesen Østergaard

**Affiliations:** Department of Clinical Medicine, Aarhus University, Aarhus, Denmark; Department of Affective Disorders, Aarhus University Hospital - Psychiatry, Aarhus, Denmark; Center for Humanities Computing, Aarhus University, Aarhus, Denmark; Deakin University, Institute for Mental and Physical Health and Clinical Translation (IMPACT), Geelong, Victoria, Australia; Mildura Base Public Hospital Mental Health Services, Alcohol and Other Drugs Integrated Treatment Team, Victoria, Australia; Barwon Health, Drugs and Alcohol Services, Mental Health Drugs and Alcohol Services, Geelong, Victoria, Australia; Psychosis Research Unit, Aarhus University Hospital - Psychiatry, Aarhus, Denmark

## Abstract

**Objective:** Electroconvulsive therapy (ECT) is an effective treatment of severe manifestations of mental illness. Since delay in initiation of ECT can have detrimental effects, prediction of the need for ECT could improve outcomes via more timely treatment initiation. This study aimed to predict the need for ECT following admission to a psychiatric hospital.

**Methods:** This cohort study was based on electronic health record (EHR) data from routine clinical practice. Adult patients admitted to a hospital within the Psychiatric Services of the Central Denmark Region between January 2013 and November 2021 were included in the study. The outcome was initiation of ECT >7 days (to not include patients admitted for planned ECT) and ≤67 days after admission. The data was randomly split into an 85% training set and a 15% test set. On the 7^th^ day of the inpatient stay, machine learning models (extreme gradient boosting (XGBoost)) were trained to predict initiation of ECT and subsequently tested on the test set, using the area under the receiver operating characteristic curve (AUROC) as the main performance measure.

**Results:** The cohort consisted of 41,610 patients with 164,961 admissions eligible for prediction. In the test set the trained model predicted ECT initiation with an AUROC of 0.94, 47% sensitivity, 98% specificity, positive predictive value of 24% and negative predictive value of 99%. The top predictors were the highest suicide assessment score and the mean Brøset violence checklist score in the preceding three months.

**Conclusions:** EHR data from routine clinical practice can be used to predict need for ECT. This may lead to more timely treatment initiation.

## Introduction

Electroconvulsive therapy (ECT) is a life-saving treatment of severe manifestations of mental illness.^1–3^ ECT is usually reserved for situations where patients have either not responded to other treatments, or when they are at imminent risk of death due to suicidality, malignant catatonia, or inanition or dehydration due to either refusal or inability to eat or drink.^3–7^ Initiation of ECT may be delayed for logistic, legal or clinical reasons (e.g., awaiting response to pharmacological treatment).^4,9,10^ This not only prolongs the burden of illness, but also seems to reduce the likelihood of response to ECT once initiated, and may even have fatal consequences.^9,11–14^ Therefore, if the need for ECT could be predicted at the level of the individual patient, there is reason to believe that it could improve outcomes via more timely treatment initiation.

There is a growing body of literature suggesting that information from electronic health records (EHR) from psychiatric services can be harnessed for clinical prediction via machine learning techniques. Indeed, we have previously demonstrated that mechanical restraint,^15^ involuntary admission,^16^ diagnostic progression to bipolar disorder or schizophrenia,^17^ type 2 diabetes,^18^ and cardiovascular disease^19^ can be predicted by machine learning models trained on EHR data, some of these outcomes with a level of prediction that bodes well for clinical implementation. Based on these experiences, the present study aimed at training and validating a machine learning model predicting the need for ECT among patients admitted to psychiatric hospitals, using only EHR data that are available from routine clinical practice.

## Methods

Reporting generally follows the Transparent Reporting of multivariable prediction models for Individual Prognosis Or Diagnosis with Artificial Intelligence (TRIPOD+AI).^20^ Figure 1 illustrates the methods used in this study, which are analogue to those used in related prediction studies based on the same data source.^15–19^

**Figure 1.**
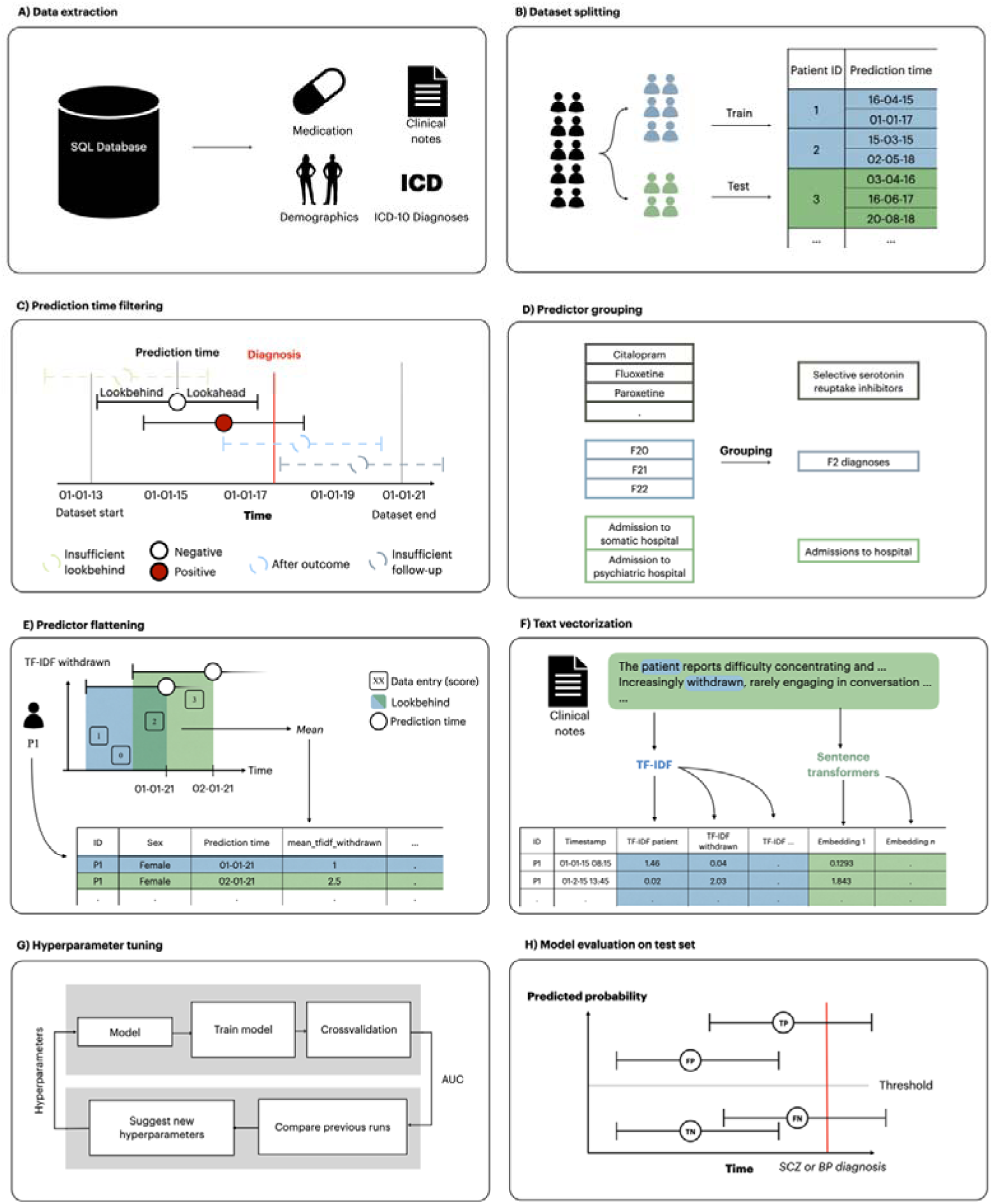
Overview of the process for extraction and transformation of dataset and the training and testing of models. A) Data were extracted from the electronic health records. B) Data were split into a training and a test set. C) Prediction times occurring after September 22, 2021 and before January 1, 2015 were removed due to lack of follow-up/lookbehind in addition to prediction times preceded by diagnoses of psychotic or personality disorders. D) Linked predictors such as medication class and diagnostic groups were grouped together. E) Predictors for each prediction time were extracted by aggregating the variables within the lookbehind with an aggregation function. As a result, each row in the dataset represents a specific prediction time with a column for each predictor. F) Clinical notes were turned into vectors using TF-IDF models. G) Models were trained and optimised on the training set using 5- fold cross-validation. Hyperparameters were tuned to optimise AUROC. H) The best candidate models were evaluated on the test set. Modified version of the figure from Bernstorff et al. 2024.^14^

### Data and Cohort Definition

The study was based on data from an updated version of the PSYchiatric Clinical Outcome Prediction (PSYCOP) cohort, encompassing routine EHR data from all individuals with at least one contact with the Psychiatric Services of the Central Denmark Region from January 1, 2011, to November 22, 2021 (Figure 1A).^21^ The Central Denmark Region has a catchment population of approximately 1.3 million. For the present study, the cohort was restricted to contacts occurring after January 1, 2013, due to inconsistencies in the data before 2013, resulting from the gradual implementation of a new electronic health record system in 2011.^22,23^ Only patients aged above 18 were included. Although often admitted for inpatient treatment, patients with a diagnosis within the schizophrenia, schizotypal and delusional disorders category (F2 in the International Statistical Classification of Diseases, Tenth Revision (ICD-10))^24^ or the disorders of adult personality and behaviour category (F6 in the ICD-10) were excluded, as ECT is primarily used for severe mood disorders in Denmark and most Western countries.^25,26^ Due to the combination of frequent admissions and rare use of ECT, including patients with prior F2 or F6 diagnoses would result in a large number of “0” values for the outcome variable, which would likely translate to poor positive predictive value of the developed models, and have limited clinical relevance.

### Data Split

The data was randomly split into an 85% training set and a 15% test set, stratified by patient. No patient appeared in both splits (Figure 1B).

### Outcome Definition

Treatment initiation with ECT (procedure codes used to identify ECT in the EHR data are available in Table S1) after a minimum of seven days of inpatient stay (to not include patients admitted for planned ECT) at a psychiatric hospital in the Central Denmark Region was used as a proxy for the need for ECT. Models were trained to predict the initiation of ECT within 60 days of the prediction time (60-day lookahead window as described below and in Figure 1C).

### Prediction Time Definition and Filtering

Prediction times were defined as the seventh day of each inpatient stay. Prediction times occurring after September 22, 2021, were removed because they did not have the required 60 days of follow-up. Predictions times occurring before January 1, 2015, were removed because they did not have sufficient data in the dataset to construct predictors using data from the 730 days prior to the prediction (Figure 1C). Furthermore, no predictions were made for patients who had received ECT within the last three years to avoid making predictions for patients already undergoing an ECT treatment course or who had a high probably of receiving a new treatment series based on relatively recent successful response to ECT.

### Predictor Construction

Only routine clinical data from the EHRs were considered for predictors, i.e., no additional data collection was performed for the purpose of this study. Predictors (some of which were grouped – see Figure 1D) from structured data were constructed by looking back a specified period (the lookbehind window) from each prediction time and extracting a single value for each predictor. When multiple values were present in the lookbehind window, an appropriate aggregation function, such as the mean or count, was applied. If no values were present in the lookbehind window, a fallback value (e.g. 0 or “not a Number” (NaN)) was used. Predictors were created using lookbehind windows of 90 days, 365 days and 730 days to incorporate different temporal contexts. Predictor construction was conducted using *timeseriesflattener v2.2.6*^27^ (Figure 1E), and included demographics (age, sex), hospital contacts (psychiatric and non-psychiatric hospital admissions, outpatient contacts with psychiatric services, psychiatric and non-psychiatric emergency visits), diagnoses, medications administered during inpatient stays, rating scales (the 17-item Hamilton Depression Rating Scale^28^ and the Brøset violence checklist^29^, suicide risk assessment (a scoring system used in the Central Denmark Region with the following risk levels: 1 - no increased risk, 2 - increased risk, and 3 - acutely increased risk)), information related to leaves during inpatient stays, and coercive measures (for the full list, see Table S2).

Free-form text from clinical notes from the EHRs (the note types are specified in Table S3 were embedded as numerical vectors using term frequency-inverse document frequency (TF-IDF) (Figure 1F). For further details, see the supplementary methods in the online supplement.

### Model Training

Separate models were trained and optimized for each of the following three predictors sets: 1) structured predictors only, 2) text predictors only, and 3) both structured and text predictors. A frequently used, state-of-the-art machine learning model, Extreme Gradient Boosting (XGBoost), was trained and hyperparameter tuned individually for each predictor set using Optuna v3.4.0 (Figure 1G).^30,31^ In prior studies based on the same dataset we have included elastic-net regularized logistic regression as a benchmark model. However, XGBoost has consistently outperformed elastic-net regularized logistic regression in these studies.^16–19^ Therefore, we chose to only train XGBoost in this study.

### Statistical Analysis

Statistical analysis was conducted from December 2022 to June 2025. Python version 3.11.8 was used for all analyses. The code for the analyses is available via GitHub.^32^ Power analyses were not conducted and sample size was determined by the available data.

### Model Evaluation

Based on the AUROC after hyperparameter tuning, the best-performing model for each predictor set (structured predictors only, text predictors only, and both structured and text predictors) was re-trained on the entire training set and applied to the test set (Figure 1H). All evaluation metrics are based on the test set unless otherwise specifically stated. The AUROC was calculated for global performance. Additionally, we report sensitivity, specificity, positive predictive value (PPV), negative predictive value (NPV), and the median time from the first positive prediction to the outcome at multiple classification thresholds. Predictor importance was estimated via information gain.

### Robustness analyses

Stratified analyses of the stability of model predictions across time since prediction, sex and age were conducted on the test set. Additionally, we assessed the generalizability of the models across both geography and time (calendar year). For geographic generalization, the model was retrained on cases from hospitals in the eastern and western parts of the region and evaluated on cases from hospitals in the central part. For temporal generalization, we retrained a series of models using data up to year *n*, incrementally including one additional year at a time, and evaluated each model on data from the following years.

### Ethics

The use of EHRs from the CDR for this study was approved by the Legal Office of the CDR in accordance with the Danish Health Care Act §46, Section 2. According to the Danish Committee Act, ethical review board approval is not required for studies based solely on data from EHRs (waiver: 1-10-72-1-22).

## Results

The cohort consisted of 41,610 unique patients (54.9% females) with a total of 164,961 inpatient stays eligible for prediction. Table 1 shows an overview of the number of patients and inpatient stays in each split, along with demographic and diagnostic characteristics. The largest predictor set contained 1,179 predictors, 179 structured predictors (listed in Table S3) and 1000 derived from clinical notes (TF-IDF).

**Table 1.** Descriptive statistics for prediction times (A) and individual patients (B) that were eligible for prediction.

|  |  | Overall | Train | Test |
| --- | --- | --- | --- | --- |
| **A. Prediction times** |  |  |  |  |
| Number of prediction times |  | 164961 | 140092 | 24869 |
| Age, median [25%,75%] |  | 52.5 [34.7,70.2] | 52.6 [34.7,70.4] | 51.8 [34.6,69.7] |
| Age grouped, n (%) | 18-19 | 2539 (1.5) | 2149 (1.5) | 390 (1.6) |
|  | 20-29 | 25614 (15.5) | 21664 (15.5) | 3950 (15.9) |
|  | 30-39 | 22006 (13.3) | 18623 (13.3) | 3383 (13.6) |
|  | 40-49 | 23050 (14.0) | 19285 (13.8) | 3765 (15.1) |
|  | 50-59 | 25834 (15.7) | 22331 (15.9) | 3503 (14.1) |
|  | 60-69 | 22048 (13.4) | 18603 (13.3) | 3445 (13.9) |
|  | 70-79 | 21859 (13.3) | 18666 (13.3) | 3193 (12.8) |
|  | 80-89 | 17600 (10.7) | 15044 (10.7) | 2556 (10.3) |
|  | 90+ | 4411 (2.7) | 3727 (2.7) | 684 (2.8) |
| Female, n (%) |  | 90629 (54.9) | 76749 (54.8) | 13880 (55.8) |
| Incident ECT within 60 days, n (%) |  | 1606 (1.0) | 1356 (1.0) | 250 (1.0) |
| *F0 - Organic disorders, n (%) |  | 17497 (10.6) | 14926 (10.7) | 2571 (10.3) |
| *F1 - Substance use disorders, n (%) |  | 27988 (17.0) | 24140 (17.2) | 3848 (15.5) |
| *F3 - Mood disorders, n (%) |  | 39984 (24.2) | 34017 (24.3) | 5967 (24.0) |
| *F4 - Anxiety disorders, n (%) |  | 34874 (21.1) | 29667 (21.2) | 5207 (20.9) |
| *F5 - Eating and sleeping disorders, n (%) |  | 3118 (1.9) | 2630 (1.9) | 488 (2.0) |
| *F7 - Mental retardation, n (%) |  | 3483 (2.1) | 2945 (2.1) | 538 (2.2) |
| *F8 - Developmental disorders, n (%) |  | 3312 (2.0) | 2917 (2.1) | 395 (1.6) |
| *F9 - Child and adolescent disorders, n (%) |  | 11508 (7.0) | 9773 (7.0) | 1735 (7.0) |
| **B. Patients** |  |  |  |  |
| Number of patients |  | 41610 | 35282 | 6328 |
| Female, n (%) |  | 24207 (58.2) | 20506 (58.1) | 3701 (58.5) |
| Incident ECT, n (%) |  | 867 (2.1) | 727 (2.1) | 140 (2.2) |

*\*These ICD-10 diagnoses (subchapters) cover the two years prior to the prediction time*

### Model training

The performance of models on the training set (based on out-of-fold data from cross-validation) indicated that those trained using either of the three predictor sets—structured, text, and the combination of the two—obtained fairly comparable results. Specifically, the model trained exclusively on structured predictors achieved an AUROC of 0.92, while the model using only text predictors attained an AUROC of 0.90. The model that integrated both structured and text predictors demonstrated marginally superior performance, with an AUROC of 0.94.

### Model testing

When applied to the test set, the models retained their performance levels observed during training (Figure 2A). Consistent with the training results, the model trained exclusively on structured predictors achieved an AUROC of 0.94, the model using only text predictors achieved an AUROC of 0.92, while the model that incorporated both structured and text predictors achieved an AUROC of 0.94. The model using only structured predictors had a substantially higher median time from the first positive prediction to ECT initiation (15.0 days) compared with the text predictor only model (9.5 days) and slightly higher time compared with the model using both structured and text predictors (13.0) days. Therefore, we show the results obtained for the model based on structured predictors in the main text, while analogous results for the two other models (text only and combined) are available in Supplement 1.

**Figure 2.**
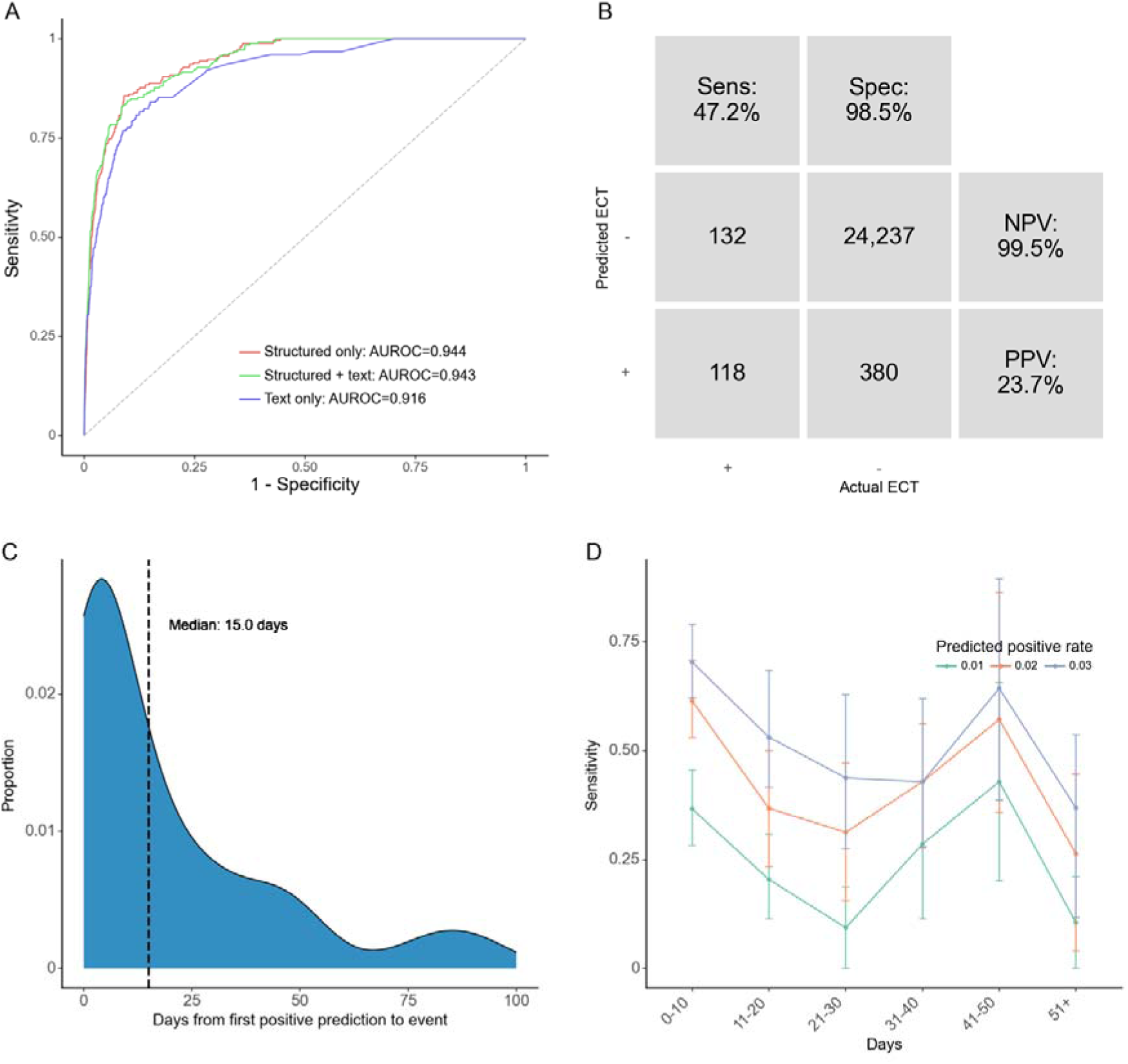
Test set results for the model based only on structured predictors. A: Receiver operating characteristics (ROC) curve for each predictor set. The model using the predictor set with only structured data was used for figures B-D with a predicted positive rate of 2%. B: Confusion matrix. PPV: Positive predictive value. NPV: Negative predictive value. C: Time (days) from the first positive prediction to the patient initiating ECT at a 2% predicted positive rate (PPR). The dashed line represents the median time. The plot is truncated at 100 days from first positive prediction to event as few events are predicted later than this. D: Sensitivity by days from prediction time to event, stratified by desired predicted positive rate (PPR).

Figure 2B shows the confusion matrix using a classification threshold based on a 2% predicted positive rate. The PPV is 23.7%, indicating that for around every 4 positive predictions, one prediction time was followed by the initiation of ECT within 60 days. The sensitivity at the level of prediction times was 47.2%, and 30.0% of all patients who received treatment with ECT were predicted positive at least once (Table S4). The median time from the first positive prediction to the outcome was 15.0 days (see Figure 2C). Notably, as evident from Figure 2C, in some cases, the model outputs a positive prediction for an admission where ECT is initiated after the first 60 days of the admission (i.e., after the end of the lookahead window). With our approach, these cases are marked as false positive predictions, even though the predictions do correctly predict ECT initiation and, thus, do hold some clinical value. As evident from Figure 2D the sensitivity of the model decreased slightly the longer into the future from the prediction time the initiation of ECT occurred.

Table 2 lists the 10 most important predictors according to information gain for the model based on structured predictors, where the suicide risk assessment score (number 1 and 4) and Brøset violence checklist score (2, 5 and 7) are particularly prominent.

**Table 2.** Top 10 most important predictors (and aggregation method) for the model based only on structured predictors.

| Predictors (aggregation function) | Information gain |
| --- | --- |
| Suicide risk assessments completed in the past 90 days (max) | 0.059 |
| Brøset violence checklist score in past 90 days (mean) | 0.056 |
| Number of temporary leaves in the past 90 days (count) | 0.053 |
| Suicide risk assessments completed in the past 90 days (most recent*) | 0.018 |
| Brøset violence checklist score in past 90 days (most recent) | 0.016 |
| Diagnosis of an organic disorder** in the past 730 days (bool) | 0.015 |
| Brøset violence checklist score in past 365 days (most recent) | 0.014 |
| Diagnosis of a substance use disorder*** in the past 730 days (bool) | 0.013 |
| Involuntary admission or detention**** in the past 90 days (max) | 0.012 |
| Physical force in the past 730 days (max) | 0.012 |

\* Most recent refers to the prior event closest to the prediction time.
\*\* ICD-10 code: F0.
\*\*\* ICD-10 code: F1.
\*\*\*\* Sectioning during an inpatient stay after being admitted voluntarily.
Clinical interpretation: Proxies for acute severity of mental illness
Clinical interpretation: Proxies for treatment resistance

### Robustness analyses

Figure 3 shows that the performance of the model based on structured predictors is stable across sexes and age groups, with slight performance reduction in older patients. Performance is relatively stable across levels of time from the first visit to the Psychiatric Services, with some instability after 25-30 months, likely partially owing to variation due to few outcomes. Lastly, no noticeable trends were observed in the performance across calendar time.

**Figure 3.**
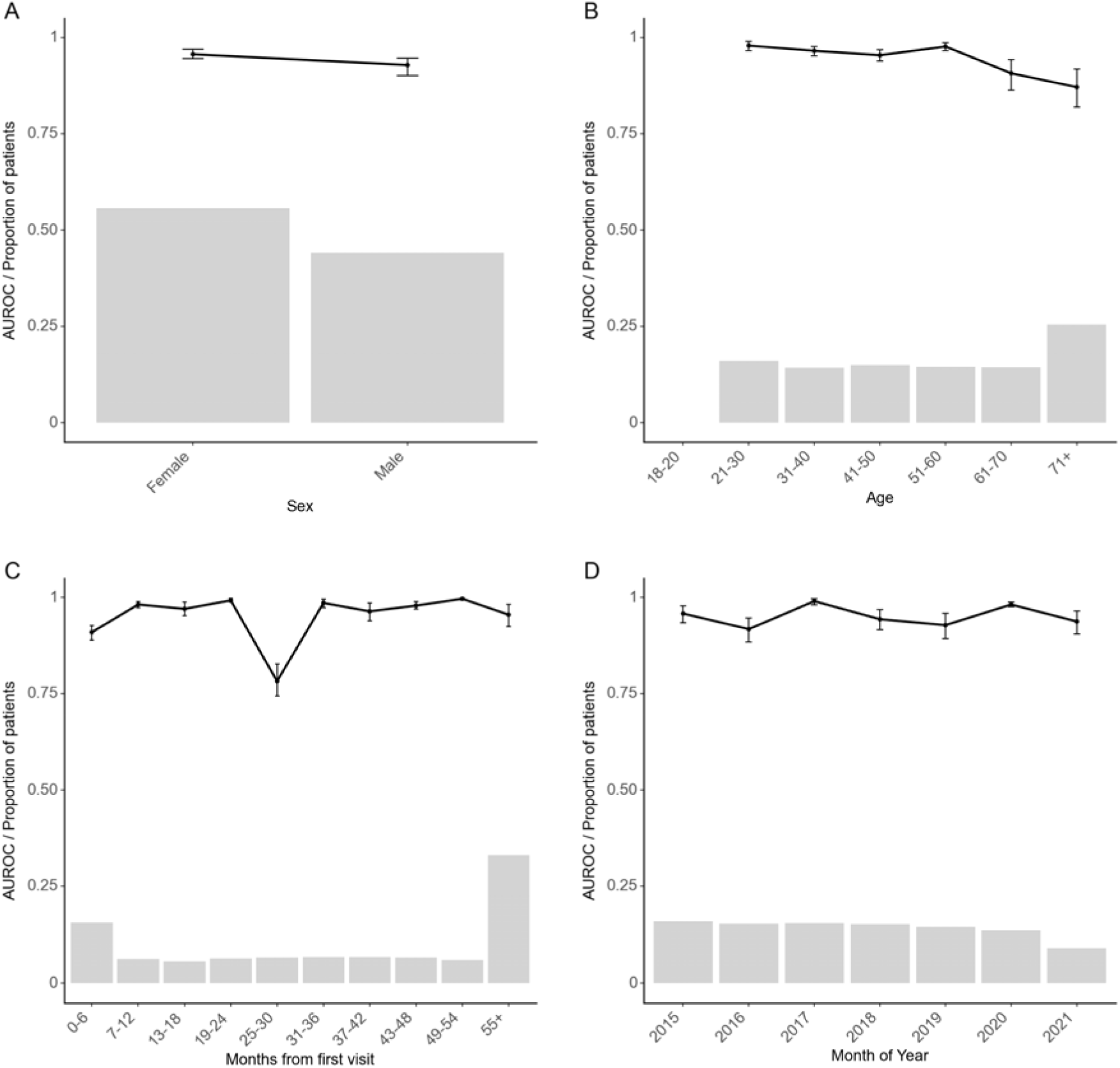
Robustness across stratifications of the model based only on structured predictors. Model performance stratified by sex (A), age in years (results for 18-20 years not reported due to too few observations) (B), time since first visit to the Psychiatric Services in the Central Denmark Region (C), and month of year (D). The black line is the area under the receiver operating characteristics curve (AUROC). Grey bars represent the proportion of prediction times that are present in each group. Error bars are 95%-confidence intervals from 100-fold bootstrap.

When evaluated on unseen hospital sites, model performance was highly similar to that obtained on unseen patients (the model using only structured predictors achieved an AUROC of 0.92). For this configuration, the median time from the first positive prediction to ECT initiation was 9.5 days, which is a substantially shorter warning time than that obtained on unseen patients (15.0 days). Finally, Figure S1 shows that the temporal stability of the model based only on structured predictors is relatively high.

### Results for models based on text predictors only and on structured + text predictors

Results analogue to those reported for the model based only on structured data in Figure 2, Table S4, Table 2 and Figure 3 and Figure S1, are reported for the model based only on text predictors (Figure S2, Table S5, Table S6, Figure S3 and Figure S4) and the model based on both structured and text predictors (Figure S5, Table S7, Table S8, Figure S6 and Figure S7) in the online supplement. When evaluated on unseen hospital sites, model performance was highly similar to that obtained on unseen patients (the model based only on text predictors achieved an AUROC of 0.91, and the model based on both structured and text predictors achieved an AUROC of 0.94). These results are generally compatible with those obtained for the model based only on structured data. The median time the first positive prediction to ECT initiation were, however, even lower for both these models (5.0 days and 6.5 days respectively).

## Discussion

The objective of this study was to investigate whether a machine learning model trained on EHR data could predict the need for ECT (proxied by initiation of ECT) among inpatients at psychiatric hospitals. Based on EHR data from 41,610 adult patients with a total of 164,961 admissions eligible for prediction, a trained machine learning model predicted ECT initiation with an area under the receiver operating characteristic curve of 0.94, obtained 47% sensitivity at a specificity of 99% with a positive predictive value of 24%, and negative predictive value of 99%. These findings suggest that it may be feasible for a machine learning prediction model to support clinical decision making and improving outcomes of ECT by allowing for more timely treatment initiation.

We are not aware of other studies having aimed at predicting the need for ECT. Therefore, there is not a literature to benchmark against. Prior machine learning studies have predominantly aimed at predicting a good response to ECT, finding that, e.g., clinical characteristics such as shorter duration of depressive episodes, depression with psychotic features, and a family history of depression may be predictive of a positive outcome of ECT.^33^ The present study addressed a different, but, we would argue, equally important, clinical problem.

Notably, the top 10 predictors driving the model seem to fall into two categories, namely i) acute severity of mental illness and ii) treatment resistance (see color coding in Table 2). With regard to the former, the fact that 2 predictors based on the suicide risk assessment score (the maximum score in the past 90 days and the most recent score in the past 90 days) were number 1, and 4 on the top 10 is in strong agreement with acute suicide risk being a prime indication for ECT.^1–3^ Similarly, three predictors based on the Brøset violence checklist (the mean score in the past 90 days, the most recent score in the past 90 days and the most recent score in the past 365 days) appeared as number 2, 5 and 7 on the top 10. Interestingly, the most recent score in the past 365 days on the 17-item Hamilton Depression Rating Scale appeared as 11^th^ most important predictor (not in the table). A high score on these scales is also indicative of acute severity of mental illness. Involuntary admission/detention and use of restraint (manual, mechanical or chemical) are, per definition, also markers of acute severity of mental illness, as this level of severity is a legal requirement for these coercive measures to be used in psychiatric services in Denmark.^34^ Finally, organic mental disorder, delirium in particular, is also strongly suggestive of acutely severe—potentially life-threatening—mental illness.^5^ Regarding treatment resistance, the number of temporary leaves (from inpatient treatment) over the past 90 days, likely reflect difficult to treatment resistant as this is suggestive of long inpatient stays with an illness severity that makes leaves possible (i.e., the patient is unlikely to be suicidal or psychotic). Finally, having received a diagnosis of substance use disorder in the past 2 years seems compatible with protracted/treatment-resistant illness.^35,36^

While the accuracy of the prediction model based on structured data is far from perfect, it may be sufficiently good to support clinical practice. If implemented, a positive prediction should ideally be used like a paraclinical test. I.e., a positive prediction should, in itself, not lead to action, but be integrated with the healthcare staff’s clinical impression of the patient, thereby contributing to an overall consideration of whether ECT may be a good treatment option in the specific case.

There are a number of limitations to consider in relation to this study. First, the word “ECT” was among the top 10 predictors in both models that included text predictors. This is likely due to these models picking up on either ECT being considered as a treatment option, being proposed to a patient, or a referral for ECT being made. This may be the prime reason why these two models had a substantially lower median time from the positive prediction to ECT initiation compared with the model based only on structured predictors, making the latter the more attractive option. Second, the models were trained exclusively to predict the need for ECT among inpatients and is unlikely to perform equally well among outpatients. In Denmark, ECT is, however, typically initiated during inpatient stays and may be continued (maintenance ECT) after discharge as part of outpatient treatment. Third, the findings from this study may not generalize to other healthcare systems with different patient populations, different use of ECT (ECT is relatively commonly used in Denmark^25,26^), and different registration practices. Fourth, we used initiation of ECT as proxy for ECT, which is not ideal as patients in need of-, but declining ECT are not covered by this proxy. Fourth, as referral to ECT is, per definition, closer in time to the need for ECT than ECT initiation, it would have been more optimal to use referrals for ECT as outcome. Unfortunately, referral data were not stable over the study period and could, therefore, not be used.

In conclusion, this study is the first to demonstrate that EHR data from routine clinical practice can be used to predict the need for ECT. These findings suggest that it may be feasible for a machine learning prediction model to support clinical decision making and improving outcomes of ECT by allowing for more timely treatment initiation.

## Author Contributions

Concept and design: Hansen, Danielsen, Østergaard

Acquisition, analysis, or interpretation of data: All authors.

Drafting of the manuscript: Hansen, Lundin, Østergaard

Critical review of the manuscript for important intellectual content: Damgaard, Danielsen. Statistical analysis: Hansen, Damgaard.

Obtained funding: Østergaard.

Supervision: Danielsen, Østergaard.

## Data availability

Due to the personally sensitive nature of the data used for this study, it cannot be shared according to Danish law.

## References

1. Salagre E, Rohde C, Østergaard SD. Self-Harm and Suicide Attempts Preceding and Following Electroconvulsive Therapy: A Population-Based Study. J ECT. 2022;38(1):13–23. doi:10.1097/YCT.0000000000000790

2. Kellner CH, Fink M, Knapp R, et al. Relief of expressed suicidal intent by ECT: a consortium for research in ECT study. Am J Psychiatry. 2005;162(5):977–982. doi:10.1176/appi.ajp.162.5.977

3. Kellner CH, Obbels J, Sienaert P. When to consider electroconvulsive therapy (ECT). Acta Psychiatr Scand. 2020;141(4):304–315. doi:10.1111/acps.13134

4. Krarup M, Kellner CH, Østergaard SD. Clinical and Legal Differences in the Use of Involuntary Electroconvulsive Therapy for Life-Threatening Illness Across European Countries. J ECT. 2024;40(2):105–110. doi:10.1097/YCT.0000000000000984

5. Salagre E, Rohde C, Ishtiak-Ahmed K, Gasse C, Østergaard SD. Survival Rate Following Involuntary Electroconvulsive Therapy: A Population-Based Study. J ECT. 2021;37(2):94–99. doi:10.1097/YCT.0000000000000736

6. Zisselman MH, Jaffe RL. ECT in the treatment of a patient with catatonia: consent and complications. Am J Psychiatry . 2010;167:127–132.

7. Liang CS, Chung CH, Ho PS, et al. Superior anti-suicidal effects of electroconvulsive therapy in unipolar disorder and bipolar depression. Bipolar Disord. 2018;20:539– 546.

8. Parry BL. The tragedy of legal impediments involved in obtaining ECT for patients unable to give informed consent. Am J Psychiatry. 1981 Aug;138(8):1128–9. doi: 10.1176/ajp.138.8.1128b. PMID: 7258400.

9. Thakrar A, Bauza P, Maaz Y, Akinyemi E. The Shocking Time It Takes to Initiate ECT: A Clinically and Legally Complicated Case of Catatonia. Prim Care Companion CNS Disord. 2018;20(6):18l-02321. Published 2018 Dec 13. doi:10.4088/PCC.18l02321

10. Livingston R, Wu C, Mu K, Coffey MJ. Regulation of Electroconvulsive Therapy: A Systematic Review of US State Laws. J ECT. 2018;34(1):60–68. doi:10.1097/YCT.0000000000000460

11. Haq, A. U., Sitzmann, A. F., Goldman, M. L., Maixner, D. F., & Mickey, B. J. (2015). Response of depression to electroconvulsive therapy: a meta-analysis of clinical predictors. The Journal of clinical psychiatry, 76(10), 18164.

12. Kindler S, Shapira B, Hadjez J, Abramowitz M, Brom D, Lerer B. (1991) Factors Influencing Response to Bilateral Electroconvulsive Therapy in Major Depression. Convulsive Therapy 7(4):p 245–254.

13. Narayanaswamy JC, Tibrewal P, Zutshi A, Srinivasaraju R, Math SB. Clinical predictors of response to treatment in catatonia. Gen Hosp Psychiatry. 2012;34(3):312–316. doi:10.1016/j.genhosppsych.2012.01.011

14. van Waarde JA, Tuerlings JH, Verwey B, van der Mast RC. Electroconvulsive therapy for catatonia: treatment characteristics and outcomes in 27 patients. J ECT. 2010;26(4):248–252. doi:10.1097/YCT.0b013e3181c18a13

15. Danielsen AA, Fenger MHJ, Østergaard SD, Nielbo KL, Mors O. Predicting mechanical restraint of psychiatric inpatients by applying machine learning on electronic health data. Acta Psychiatr Scand. 2019;140(2):147–157. doi:10.1111/acps.13061

16. Perfalk E, Damgaard JG, Bernstorff M, Hansen L, Danielsen AA, Østergaard SD. Predicting involuntary admission following inpatient psychiatric treatment using machine learning trained on electronic health record data. Psychol Med. Published online November 18, 2024. doi:10.1017/S0033291724002642

17. Hansen L, Bernstorff M, Enevoldsen K, et al. Predicting Diagnostic Progression to Schizophrenia or Bipolar Disorder via Machine Learning. JAMA Psychiatry. Published online February 19, 2025. doi:10.1001/jamapsychiatry.2024.4702

18. Bernstorff M, Hansen L, Enevoldsen K, et al. Development and validation of a machine learning model for prediction of type 2 diabetes in patients with mental illness. Acta Psychiatr Scand. 2025;151(3):245–258. doi:10.1111/acps.13687

19. Bernstorff M, Hansen L, Olesen KKW, Danielsen AA, Østergaard SD. Predicting cardiovascular disease in patients with mental illness using machine learning. Eur Psychiatry. 2025;68(1):e12. Published 2025 Jan 8. doi:10.1192/j.eurpsy.2025.1

20. Collins GS, Moons KGM, Dhiman P, et al. TRIPOD+AI statement: updated guidance for reporting clinical prediction models that use regression or machine learning methods. BMJ. 2024;385:e078378. doi:10.1136/bmj-2023-078378

21. Hansen L, Enevoldsen KC, Bernstorff M, Nielbo KL, Danielsen AA, Østergaard SD. The PSYchiatric clinical outcome prediction (PSYCOP) cohort: leveraging the potential of electronic health records in the treatment of mental disorders. Acta Neuropsychiatr. 2021;33(6):323–330. doi:10.1017/neu.2021.22

22. Bernstorff M, Hansen L, Perfalk E, Danielsen AA, Østergaard SD. Stability of diagnostic coding of psychiatric outpatient visits across the transition from the second to the third version of the Danish National Patient Registry. Acta Psychiatr Scand. 2022;146(3):272–283. doi:10.1111/acps.13463

23. Hansen L, Enevoldsen K, Bernstorff M, et al. Lexical stability of psychiatric clinical notes from electronic health records over a decade. Acta Neuropsychiatr. Published online August 25, 2023:1–11. doi:10.1017/neu.2023.46

24. World Health Organization. The ICD-10 Classification of Mental and Behavioural Disorders: Diagnostic Criteria for Research. World Health Organization; 1993.

25. Leiknes KA, Jarosh-von Schweder L, Høie B. (2012) Contemporary use and practice of electroconvulsive therapy worldwide. Brain Behav. 2(3):283–344. doi: 10.1002/brb3.37. PMID: 22741102; PMCID: PMC3381633.

26. Bjørnshauge D, Hjerrild S, Videbech P. Electroconvulsive Therapy Practice in the Kingdom of Denmark: A Nationwide Register- and Questionnaire-Based Study. J ECT. 2019;35(4):258–263. doi:10.1097/YCT.0000000000000586

27. Bernstorff M, Enevoldsen K, Damgaard J, Danielsen A, Hansen L. timeseriesflattener: A Python package for summarizing features from (medical) time series. J Open Source Softw. 2023;8(83):5197. doi:10.21105/joss.05197

28. Hamilton M. A rating scale for depression. J Neurol Neurosurg Psychiatry. 1960;23(1):56.

29. Woods P, Almvik R. The Brøset violence checklist (BVC). Acta Psychiatr Scand Suppl. 2002;(412):103–105. doi:10.1034/j.1600-0447.106.s412.22.x

30. Akiba T, Sano S, Yanase T, Ohta T, Koyama M. Optuna: A Next-generation Hyperparameter Optimization Framework. In: Proceedings of the 25th ACM SIGKDD International Conference on Knowledge Discovery & Data Mining. KDD ’19. Association for Computing Machinery; 2019:2623–2631. doi:10.1145/3292500.3330701

31. Chen T, Guestrin C. XGBoost: A Scalable Tree Boosting System. In: Proceedings of the 22nd ACM SIGKDD International Conference on Knowledge Discovery and Data Mining. KDD ’16. Association for Computing Machinery; 2016:785–794. doi:10.1145/2939672.2939785

32. Aarhus Psychiatry Research. Github. Updated March 2025. https://github.com/Aarhus-Psychiatry-Research Last accessed: March 1, 2025.

33. Lundin RM, Falcao VP, Kannangara S, et al. Machine Learning in Electroconvulsive Therapy: A Systematic Review. J ECT. Published online June 10, 2024. doi:10.1097/YCT.000000000000100930.

34. Danish Laws. Psykiatriloven (The Danish Mental Health Act): https://danskelove.dk/psykiatriloven Last accessed: March 7, 2025).

35. Brenner P, Brandt L, Li G, DiBernardo A, Bodén R, Reutfors J. Treatment- resistant depression as risk factor for substance use disorders-a nation-wide register-based cohort study. Addiction. 2019;114(7):1274–1282.

36. Lundberg J, Cars T, Lööv SÅ, Söderling J, Sundström J, Tiihonen J, Leval A, Gannedahl A, Björkholm C, Själin M, Hellner C. Association of Treatment- Resistant Depression With Patient Outcomes and Health Care Resource Utilization in a Population-Wide Study. JAMA Psychiatry. 2023;80(2):167–175.

