## Supplementary material for "Predicting the need for electroconvulsive therapy via machine learning trained on electronic health record data": Online Supplement

Supplementary Methods

In TF-IDF, each clinical note is represented as a list of numbers, where each number corresponds to unique terms (words). The value of each number represents the frequency of the word within the specific note divided by the frequency of the word across all notes. This results in a list of numbers which emphasizes (i.e., high values) words that are distinctive of the note in question. TF-IDF is widely used due to its interpretability/explainability and high performance both in terms of speed of computation and quality of results. Accordingly, our prior use of TF-IDF has yielded results with high face validity.^1,2^ Models were trained on all clinical notes specified in eTable 3 using *scikit-learn* v1.2.1^3^ with a dimensionality of 1000 predictors. The text embeddings were processed similarly to the structured predictors, but only a single lookbehind (730 days) and the mean aggregation function was used

**Table S1.** Procedure codes used to identify electroconvulsive therapy (ECT) in the electronic health records

BRXA1 - Voluntary treatment with ECT
BRXA10 - Voluntary treatment with unilateral ECT
BRXA11 - Voluntary treatment with bilateral ECT

BRTB1 - Involuntary treatment with ECT
BRTB10 - Involuntary treatment with unilateral ECT
BRTB11 - Involuntary treatment with bilateral ECT

BRTB10A - Involuntary treatment with unilateral ECT due to life-threatening condition
BRTB11A - Involuntary treatment with bilateral ECT due to life-threatening condition
BRTB11B - Involuntary treatment with bilateral ECT due to danger*

* There were very few ECT treatments registered with BRTB11B in our total dataset (6 out of more than 55,000) and they are probably all mislabellings of BRTB11A, as life-threatening condition is the only situation in which involuntary ECT can be used according to the Danish Mental Health Act.

**Table S2.** Overview of the 161 structured predictors used for the main model

|  | | **Group** | **Data variable** | **Lookbehind (days)** | **Aggregation** | **N** |
| --- | --- | --- | --- | --- | --- | --- |
|  | | **Static** | Sex (male/female)  Age (years) | - | - | 2 |
|  | | **Hospital contacts** | Outpatient visits  Psychiatric | 90, 365, 730 | Count, sum of hours | 6 |
|  | |  | Admissions  Psychiatric  Non-psychiatric | 90, 365, 730 | Count, sum of hours | 12 |
|  | |  | Emergency visits  Psychiatric  Non-psychiatric | 90, 365, 730 | Count | 6 |
|  |  | | All physical visits  Psychiatric  Non-psychiatric | 90, 365, 730 | Count | 6 |
|  | | **Diagnoses (ICD-10)** | F0: Organic mental disorders  F1: Mental and behavioural disorders due to psychoactive substance use  F2: Schizophrenia, schizotypal, and delusional disorders  F20: Schizophrenia  F25: Schizoaffective  F3: Mood/affective disorders  F30-F31: Manic and bipolar disorders  F4: Neurotic, stress-related, and somatoform disorders  F5: Behavioural syndromes associated with physiological disturbances and physical factors  F6: Disorders of adult personality and behaviour  F60.2-60.4 Cluster B personality (dissocial-, borderline- and histrionic personality disorder)  F7: Mental retardation  F8: Disorders of psychological development  F9: Behavioural and emotional disorders with onset usually occurring in childhood and adolescence or unspecified mental disorder | 90, 365, 730 | Boolean | 30 |
|  | | **Medication** | Antipsychotics  1.generation  2. Generation  Clozapine  Benzodiazepines  Benzodiazepine related sleeping agents  Lamotrigine  Lithium  NaSSAs  Pregabaline  SNIRs  SRRIs TCAs  Valproate | 90, 365, 730 | Boolean | 39 |
|  | |  | All antipsychotics  All antidepressants | 90, 365, 730 | Count | 6 |
|  | | **Coercive measures** | Detention or involuntary admission Involuntary treatment  Restraint (manual, mechanical and chemical) | 90, 365, 730 | Latest, max, mean | 27 |
|  | | **Psychometric rating scales** | Brøset Violence Checklist score  Hamilton-D17 score | 90, 365, 730 | Latest, mean | 12 |
|  | |  | Suicide Risk Assessment | 90, 365, 730 | Latest, max, mean | 9 |
| **Leave** | | | No temporary leave Supervised temporary leave Unsupervised temporary leave Any temporary leave | 90, 365, 730 | Boolean, count | 24 |

**Table S3.** Clinical note types included in the generation of text predictors*

| **Danish name** | **English name** | **Description** |
| --- | --- | --- |
| Aftaler, Psykiatri | Appointments, Psychiatry | Description of care- and treatment-related appointments and agreements with and about the patient. |
| Aktuelt socialt, Psykiatri | Current social functioning | Description of the patient’s current social situation: relationship to the family, civil status, residential-, occupational-, and economic conditions, and contact with the social services. |
| Aktuelt psykisk | Subjective mental state | Description of the development, progress, and current status of the patient’s mental illness incl. description of symptoms. |
| Aktuelt somatisk, Psykiatri | Subjective physical state | Description of the patient’s current and chronic somatic illnesses and symptoms. Information on current treatment in relation to the patient’s physical condition. |
| Konklusion/vurdering | Conclusion/evaluation | Aggregation and interpretation of all findings and decisions based on an interview (e.g., in relation to an outpatient visit or during an inpatient stay). |
| Kontaktårsag | Reason of contact | The reason for the patient’s in- or outpatient treatment course. |
| Objektivt psykisk | Current objective mental state | Objective assessment of the patient’s mental state, including state of consciousness, orientation, intelligence, psychomotor function, mood, delusions, psychotic- and other symptoms, etc. |
| Observation af patient, Psykiati | Observation of patient, Psychiatry | Description of an inpatient’s symptoms, behaviour, reactions towards relatives, other patients, staff, etc. |
| Samtale med behandlingssigte | Conversation with treatment aim | Documentation of the conversation’s purpose and attendees. |
| Semistruktureret diagnostisk interview | Semi-structured diagnostic interview | Registration that a semi-structured diagnostic interview has been conducted. Description of the method and documentation of result and conclusion. |
| Telefonnotat | Telephone note | Documentation of telephone conversations with a patient or the patient’s guardian. |

 *Modified version of eTable 2 from Hansen et al.^1^

### Table S4. Performance by predicted positive rate on the test set for the model based only on structured predictors

| ***Predicted positive rate*** | ***True prevalence*** | ***PPV*** | ***NPV*** | ***FPR*** | ***FNR*** | ***Sens*** | ***Spec*** | ***Acc*** | ***TP*** | ***TN*** | ***FP*** | ***FN*** | ***% of all ECT captured*** | ***F1*** | ***Median days from first positive to ECT*** |
| --- | --- | --- | --- | --- | --- | --- | --- | --- | --- | --- | --- | --- | --- | --- | --- |
| *1.0%* | *1.0%* | *27.3%* | *99.3%* | *0.7%* | *72.8%* | *27.2%* | *99.3%* | *98.5%* | *68* | *24,436* | *181* | *182* | *19.3%* | *27.3%* | *14* |
| *2.0%* | *1.0%* | *23.7%* | *99.5%* | *1.5%* | *52.8%* | *47.2%* | *98.5%* | *97.9%* | *118* | *24,237* | *380* | *132* | *30.0%* | *31.6%* | *15* |
| *3.0%* | *1.0%* | *19.0%* | *99.6%* | *2.5%* | *43.2%* | *56.8%* | *97.5%* | *97.1%* | *142* | *24,013* | *604* | *108* | *36.4%* | *28.5%* | *14* |
| *4.0%* | *1.0%* | *16.3%* | *99.6%* | *3.4%* | *35.2%* | *64.8%* | *96.6%* | *96.3%* | *162* | *23,784* | *833* | *88* | *44.3%* | *26.0%* | *16* |

*Predicted positive rate: The proportion of contacts predicted positive by the model. Since the model outputs a predicted probability, this is a threshold set by us. True prevalence: The proportion of contacts that received treatment with ECT within the lookahead window. PPV: Positive predictive value. NPV: Negative predictive value. FPR: False positive rate. FNR: False negative rate. TP: True positives. Numbers are service contacts. TN: True negatives. Numbers are service contacts. FP: False positives. Numbers are service contacts. FN: False negatives. Numbers are service contacts. % of all ECT captured: Percentage of all patients who received ECT, who had at least one positive prediction. Median days from first positive to ECT: For all patients with at least one true positive prediction, the number of days from their first positive prediction to initiating ECT.*

**Figure S1.** Temporal stability of the model based only on structured predictors

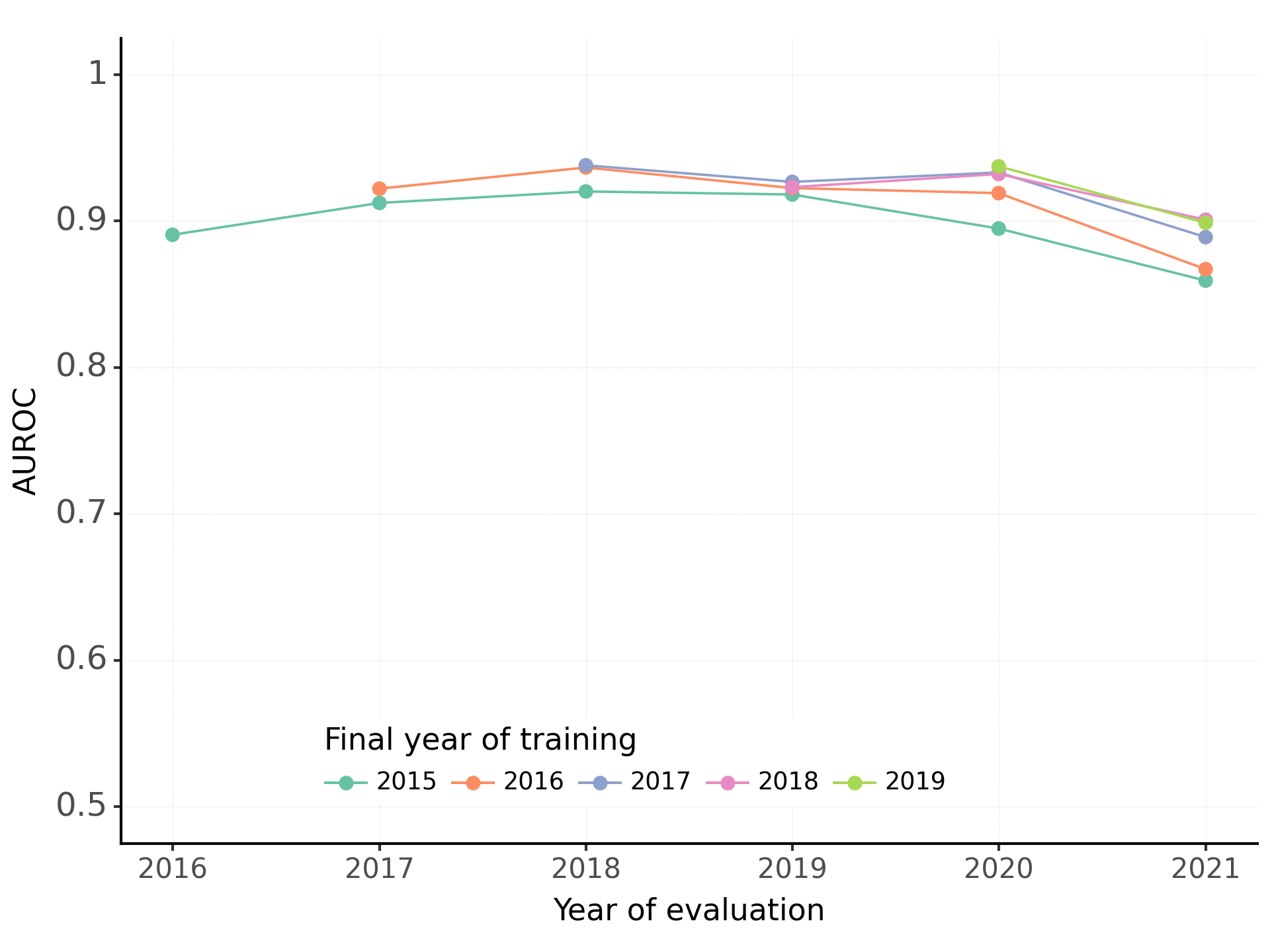

**Figure S2.** Test set results for the model based only on text predictors

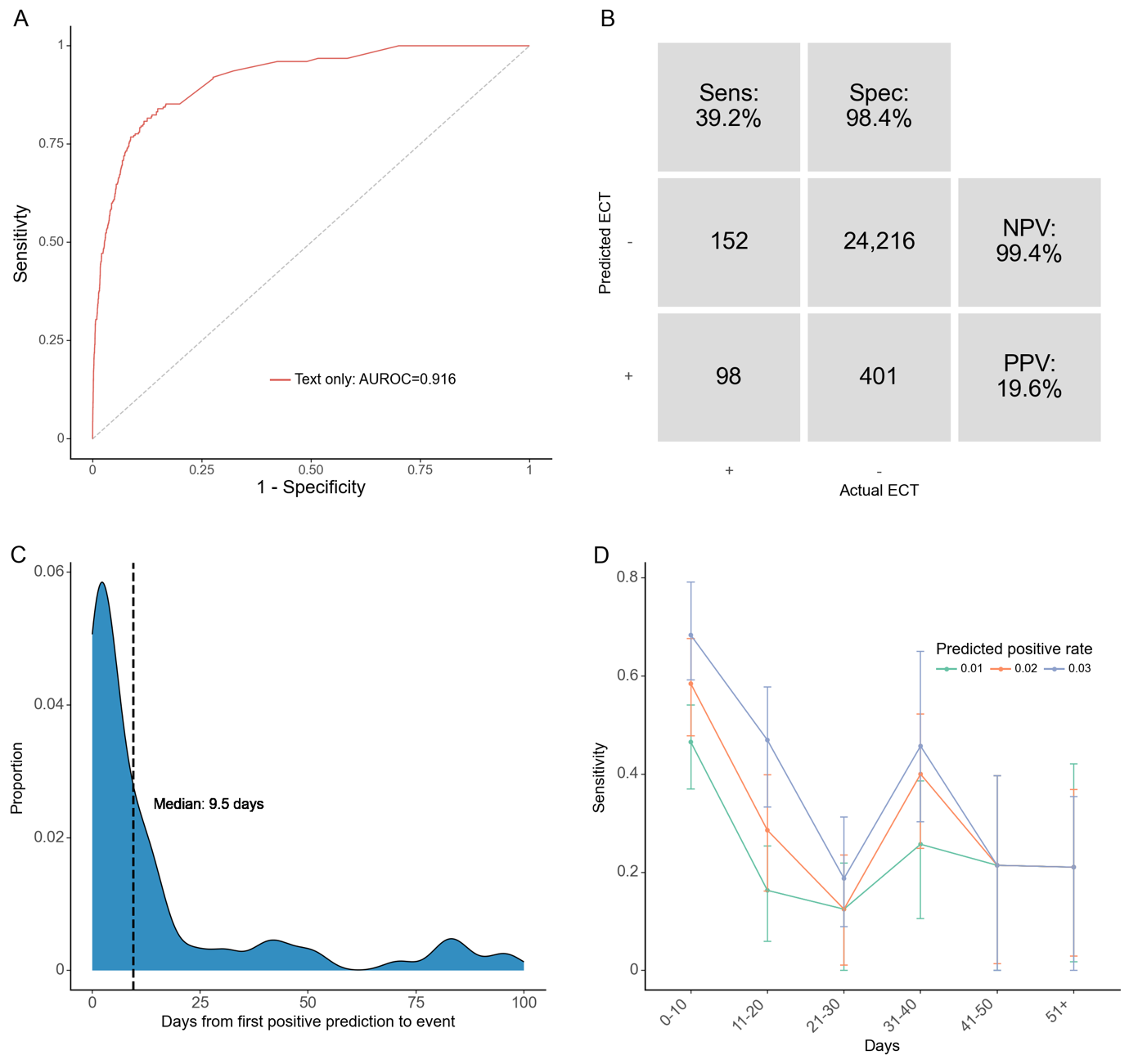

*A: Receiver operating characteristics (ROC) curve for each predictor set. The model using the predictor set with only text data was used for figures B-D with a predicted positive rate of 2%. B: Confusion matrix. PPV: Positive predictive value. NPV: Negative predictive value. C: Time (days) from the first positive prediction to the patient receiving treatment with ECT at a 2% predicted positive rate (PPR). The dashed line represents the median time. The plot is truncated at 100 days from first positive prediction to event as few events are predicted later than this. D: Sensitivity by days from prediction time to event, stratified by desired predicted positive rate (PPR).*

**Table S5.** Performance by predicted positive rate on the test set for the model based only on text predictors

| ***Predicted positive rate*** | ***True prevalence*** | ***PPV*** | ***NPV*** | ***FPR*** | ***FNR*** | ***Sens*** | ***Spec*** | ***Acc*** | ***TP*** | ***TN*** | ***FP*** | ***FN*** | ***% of all ECT captured*** | ***F1*** | ***Median days from first positive to ECT*** |
| --- | --- | --- | --- | --- | --- | --- | --- | --- | --- | --- | --- | --- | --- | --- | --- |
| *1.0%* | *1.0%* | *30.1%* | *99.3%* | *0.7%* | *70.0%* | *30.0%* | *99.3%* | *98.6%* | *75* | *24,443* | *174* | *175* | *20.7%* | *30.1%* | *7,5* |
| *2.0%* | *1.0%* | *19.6%* | *99.4%* | *1.6%* | *60.8%* | *39.2%* | *98.4%* | *97.8%* | *98* | *24,216* | *401* | *152* | *24.3%* | *26.2%* | *9,5* |
| *3.0%* | *1.0%* | *16.2%* | *99.5%* | *2.5%* | *51.6%* | *48.4%* | *97.5%* | *97.0%* | *121* | *23,992* | *625* | *129* | *27.9%* | *24.3%* | *12* |
| *4.0%* | *1.0%* | *13.7%* | *99.5%* | *3.5%* | *45.6%* | *54.4%* | *96.5%* | *96.1%* | *136* | *23,758* | *859* | *114* | *32.1%* | *21.8%* | *14* |

*Predicted positive rate: The proportion of contacts predicted positive by the model. Since the model outputs a predicted probability, this is a threshold set by us. True prevalence: The proportion of contacts that received treatment with ECT within the lookahead window. PPV: Positive predictive value. NPV: Negative predictive value. FPR: False positive rate. FNR: False negative rate. TP: True positives. Numbers are service contacts. TN: True negatives. Numbers are service contacts. FP: False positives. Numbers are service contacts. FN: False negatives. Numbers are service contacts. % of all ECT captured: Percentage of all patients who received ECT, who had at least one positive prediction. Median days from first positive to ECT: For all patients with at least one true positive prediction, the number of days from their first positive prediction to initiating ECT.*

**Table S6.** Top 10 most important predictors for the model based only on text predictors

| **Predictors* (Danish)** | **English translation** | **Information gain** |
| --- | --- | --- |
| Stue | Patient room | 0.017 |
| Stuen | The patient room | 0.010 |
| Besøg | Visit | 0.007 |
| ECT | ECT | 0.005 |
| Indlagt | Admitted | 0.005 |
| Mimik | Facial expression | 0.004 |
| Vagten | The shift (as in nightshift) | 0.004 |
| Oxapax | Trade name for Oxazepam | 0.003 |
| Grad | Degree (as in severity degree) | 0.003 |
| Indlæggelse | Admission | 0.003 |

* All predictors are text frequency-inverse document frequency (TF-IDF) with a 2-year look-behind and mean aggregation function.

**Figure S3.** Robustness across stratifications of the model based only on text predictors

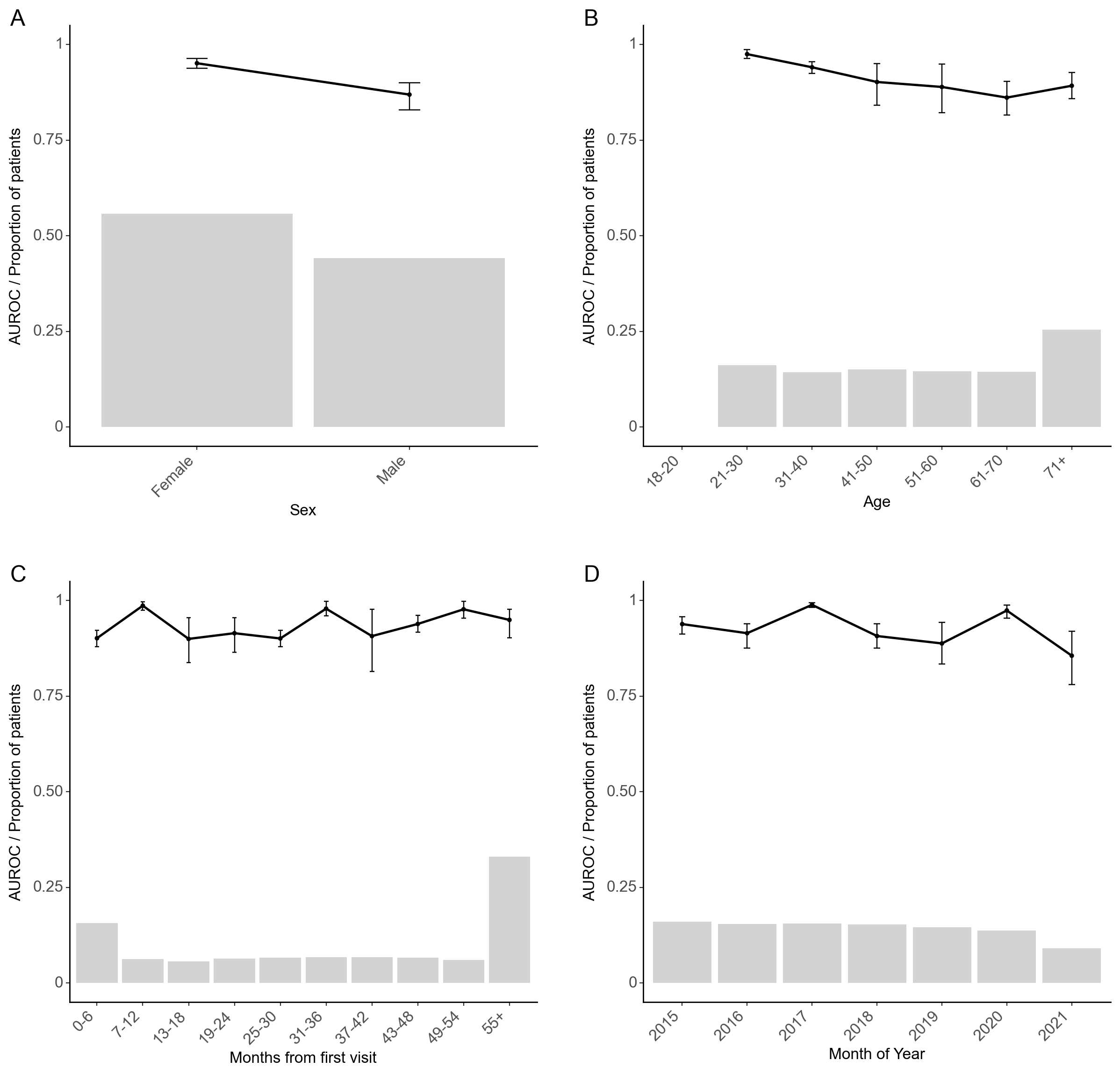

*Model performance stratified by sex (A), age in years (results for 18-20 years not reported due to too few observations) (B), time since first visit to the Psychiatric Services in the Central Denmark Region (C), and month of year (D). The black line is the area under the receiver operating characteristics curve. Grey bars represent the proportion of prediction times that are present in each group. Error bars are 95%-confidence intervals from 100-fold bootstrap.*

**Figure S4.** Temporal stability of the model based only on text predictors

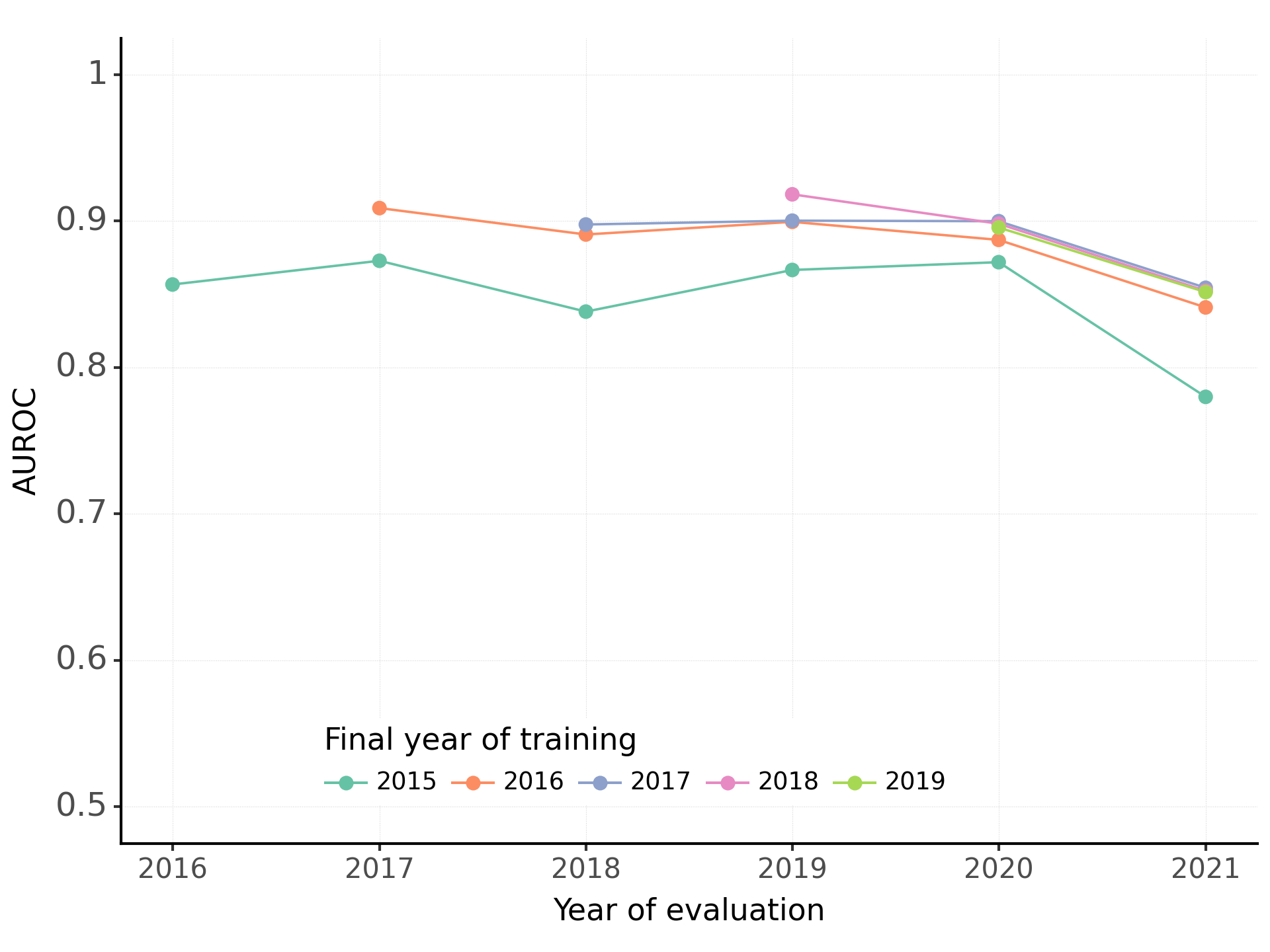

### Figure S5. Test set results for the model based on both structured and text predictors

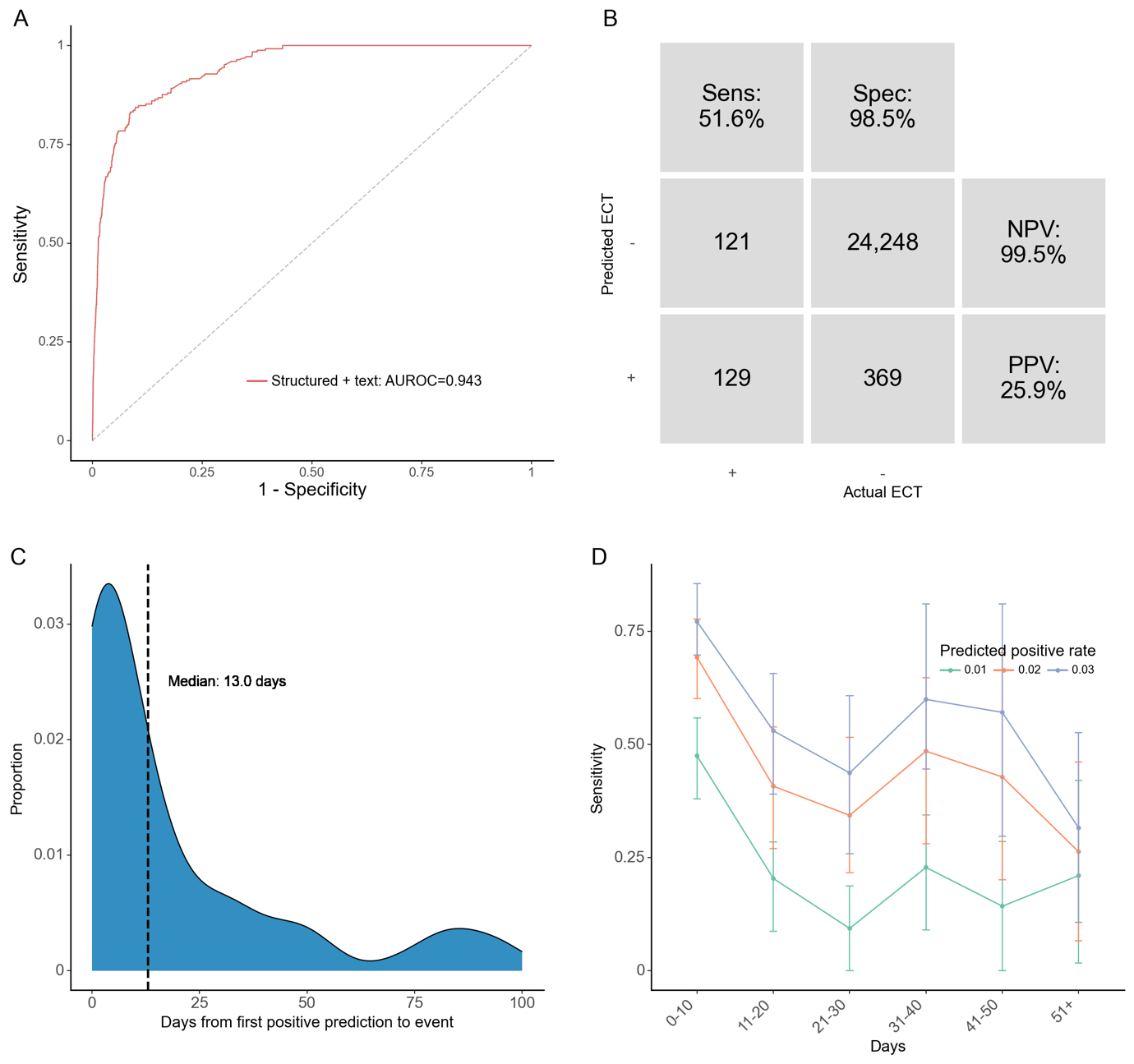

*A: Receiver operating characteristics (ROC) curve for each predictor set. The model using the predictor set with the highest AUROC (structured + text) was used for figures B-D with a predicted positive rate of 2%. B: Confusion matrix. PPV: Positive predictive value. NPV: Negative predictive value. C: Time (days) from the first positive prediction to the patient receiving treatment with ECT at a 2% predicted positive rate (PPR). The dashed line represents the median time. The plot is truncated at 100 days from first positive prediction to event as few events are predicted later than this. D: Sensitivity by days from prediction time to event, stratified by desired predicted positive rate (PPR).*

**Table S7.** Performance by predicted positive rate on the test set for the model based on both structured and text predictors

| ***Predicted positive rate*** | ***True prevalence*** | ***PPV*** | ***NPV*** | ***FPR*** | ***FNR*** | ***Sens*** | ***Spec*** | ***Acc*** | ***TP*** | ***TN*** | ***FP*** | ***FN*** | ***% of all ECT captured*** | ***F1*** | ***Median days from first positive to ECT*** |
| --- | --- | --- | --- | --- | --- | --- | --- | --- | --- | --- | --- | --- | --- | --- | --- |
| *1.0%* | *1.0%* | *30.1%* | *99.3%* | *0.7%* | *70.0%* | *30.0%* | *99.3%* | *98.6%* | *75* | *24,443* | *174* | *175* | *19.3%* | *30.1%* | *3,5* |
| *2.0%* | *1.0%* | *25.9%* | *99.5%* | *1.5%* | *48.4%* | *51.6%* | *98.5%* | *98.0%* | *129* | *24,248* | *369* | *121* | *36.4%* | *34.5%* | *13* |
| *3.0%* | *1.0%* | *20.5%* | *99.6%* | *2.4%* | *38.8%* | *61.2%* | *97.6%* | *97.2%* | *153* | *24,022* | *595* | *97* | *40.0%* | *30.7%* | *15,5* |
| *4.0%* | *1.0%* | *16.8%* | *99.7%* | *3.4%* | *33.2%* | *66.8%* | *96.6%* | *96.3%* | *167* | *23,789* | *828* | *83* | *42.9%* | *26.8%* | *18* |

*Predicted positive rate: The proportion of contacts predicted positive by the model. Since the model outputs a predicted probability, this is a threshold set by us. True prevalence: The proportion of contacts that received treatment with ECT within the lookahead window. PPV: Positive predictive value. NPV: Negative predictive value. FPR: False positive rate. FNR: False negative rate. TP: True positives. Numbers are service contacts. TN: True negatives. Numbers are service contacts. FP: False positives. Numbers are service contacts. FN: False negatives. Numbers are service contacts. % of all ECT captured: Percentage of all patients who received ECT, who had at least one positive prediction. Median days from first positive to ECT: For all patients with at least one true positive prediction, the number of days from their first positive prediction to initiating ECT.*

**Figure S6.** Robustness across stratifications of the model based on both structured and text predictors

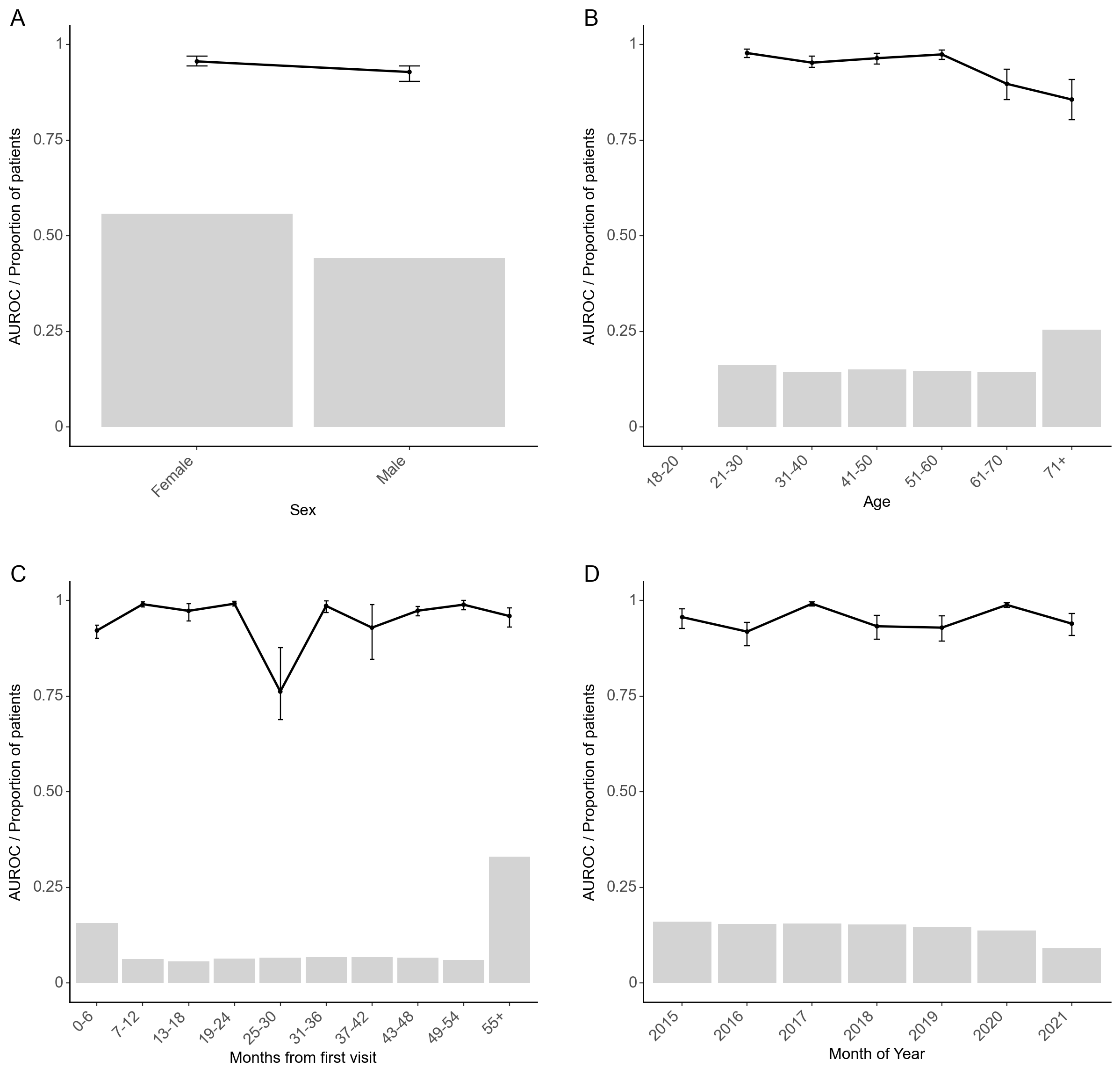

*Model performance stratified by sex (A), age in years (results for 18-20 years not reported due to too few observations) (B), time since first visit to the Psychiatric Services in the Central Denmark Region (C), and month of year (D). The black line is the area under the receiver operating characteristics curve. Grey bars represent the proportion of prediction times that are present in each group. Error bars are 95%-confidence intervals from 100-fold bootstrap.*

**Table S8.** Top 10 most important predictors for the model based on both structured and text predictors.

| **Predictors (aggregation function)** | **English translation** | **Information gain** |
| --- | --- | --- |
| Broeset violence checklist 90-day (mean) |  | 0.014 |
| Physical visits to psychiatry 90-day (count) | The patient room | 0.012 |
| Suicide risk assessment 90-day (max) |  | 0.011 |
| Temporary leave 90-day (count) |  | 0.007 |
| Substance use disorder 730-day (bool) |  | 0.007 |
| Broeset violence checklist 90-day (latest) |  | 0.007 |
| TF-IDF* “ECT” | ECT | 0.007 |
| Mood/affective disorder 90-day (bool) |  | 0.005 |
| TF-IDF* “Depression” | Depression | 0.005 |
| Suicide risk assessment 90-day (latest) |  | 0.005 |

* The 3 TF-IDF (text frequency-inverse document frequency) predictors are all based on a 2-year look-behind and the mean aggregation function.

** Noradrenergic and specific serotonergic antidepressants

**Figure S7.** Temporal stability of the model based on both structured and text predictors

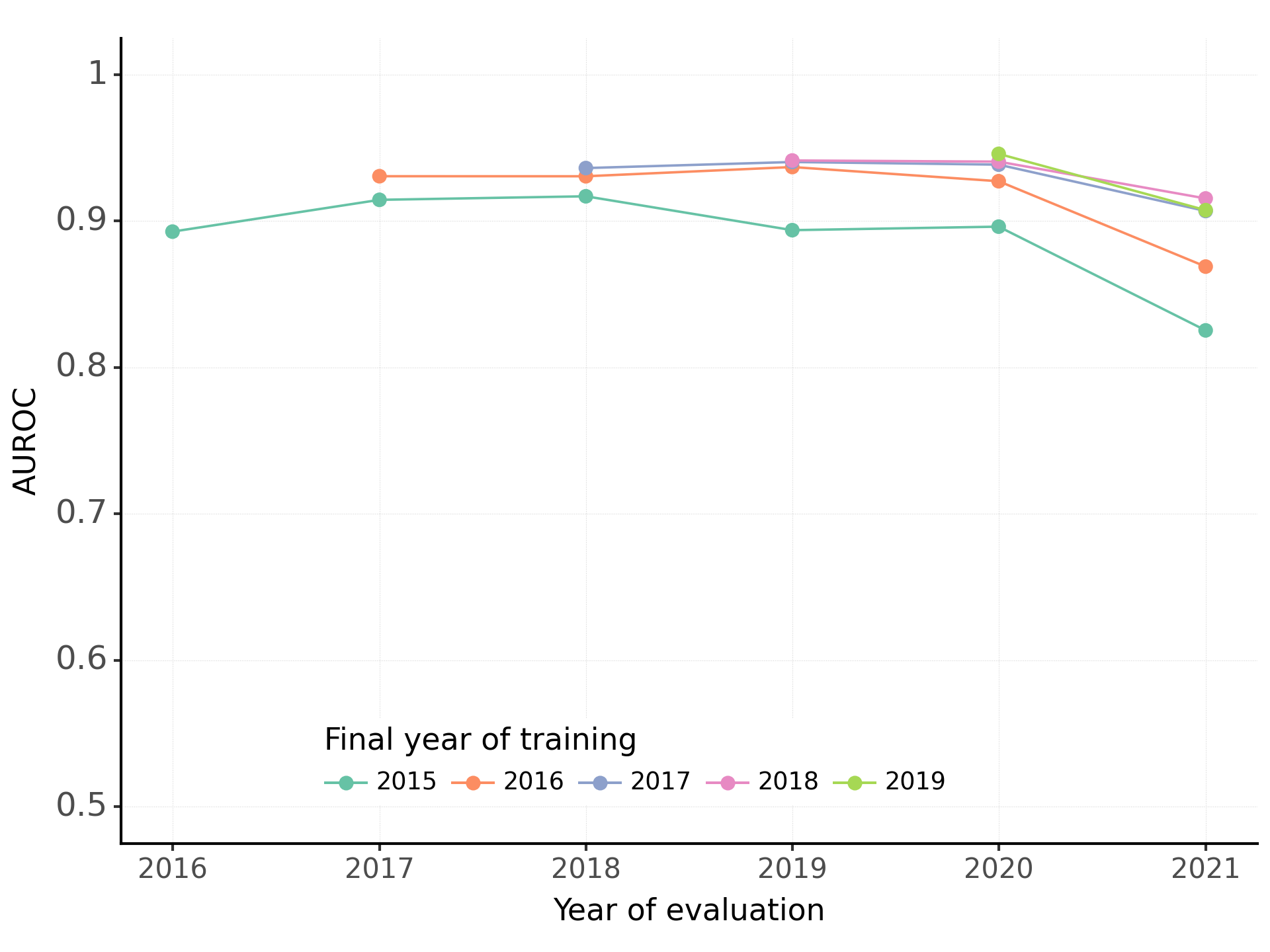

References

1. Hansen L, Bernstorff M, Enevoldsen K, et al. Predicting Diagnostic Progression to Schizophrenia or Bipolar Disorder via Machine Learning. *JAMA Psychiatry*. Published online February 19, 2025. doi:10.1001/jamapsychiatry.2024.4702
2. Perfalk E, Damgaard JG, Bernstorff M, Hansen L, Danielsen AA, Østergaard SD. Predicting involuntary admission following inpatient psychiatric treatment using machine learning trained on electronic health record data. *Psychol Med*. Published online November 18, 2024. doi:10.1017/S0033291724002642
3. Pedregosa F, Varoquaux G, Gramfort A, et al. Scikit-learn: Machine learning in Python. *J Mach Learn Res*. 2011;12:2825-2830.
